# Normal Tissue Complication Probability (NTCP) prediction model for osteoradionecrosis of the mandible in head and neck cancer patients following radiotherapy: Large-scale observational cohort

**DOI:** 10.1101/2021.03.04.21252505

**Authors:** Lisanne V. van Dijk, Abdelrahman A. Abusaif, Jillian Rigert, Mohamed A. Naser, Katherine A. Hutcheson, Stephen Y. Lai, Clifton D. Fuller, Abdallah S. R. Mohamed, on behalf on the MD Anderson Symptom Working Group

## Abstract

**Background and purpose:** Osteoradionecrosis (ORN) of the mandible represents a severe, debilitating complication of radiation therapy (RT) for head and neck cancer (HNC). At present, no Normal Tissue Complication Probability (NTCP) models for risk of ORN exist. The aim of this study was to develop a multivariable clinical/dose-based NTCP model for the prediction of ORN any grade (ORN_I-IV_) and Grade IV (ORN_IV_) following radiotherapy (± chemotherapy) in HNC patients.

**Methods:** Included HNC patients were treated with (chemo-)radiotherapy between 2005 and 2015. Mandible bone radiation dose-volume parameters, and clinical variables (i.e. *age, sex, tumor site, pre-RT dental extractions, chemotherapy history, post-operative RT* and *smoking status)*, were considered as potential predictors. The patient cohort was randomly divided into a training (70%) and independent test (30%) cohort. Bootstrapped forward variable selection was performed in the training cohort to select the predictors for the NTCP models. Final NTCP model(s) were validated on the holdback test subset.

**Results:** Of 1259 included HNC patients, 13.7% (n=173 patients) developed any grade ORN (ORNI_I-IV_ primary endpoint) and 5% (n=65) ORN_IV_ (secondary endpoint). All dose and volume parameters of the mandible bone were significantly associated with the development of ORN in univariable models. Multivariable analyses identified D_30%_ and pre-RT *dental extraction* as independent predictors for both ORN_I-IV_ and ORN_IV_ best-performing NTCP models with an AUC of 0.78 (AUC_validation_=0.75) and 0.81 (AUC_validation_=0.82), respectively.

**Conclusion:** This study presented NTCP models – based on mandible bone D_30%_ and *pre-RT dental extraction* – that predict ORN_I-IV_ and ORN_IV_ (i.e. needing invasive surgical intervention) following HNC radiotherapy. Our results suggest that less than 30% of the mandible should receive a dose of 35Gy or more for an ORN_I-IV_ risk lower than 5%. These NTCP models can improve ORN prevention and management by identifying patients at risk of ORN.

## Introduction

Osteoradionecrosis (ORN) of the mandible is a severe late toxicity following chemo-radiation for head and neck cancer (HNC) with a reported incidence between 1-16% [1–4]. While ORN is less prevalent relative to other radiation-attributable HNC toxicities, ORN is often extremely debilitating, require intense resource requirements for management, and contributes to a substantial negative impact on the quality of life [5]. With the rising incidence of human papillomavirus (HPV) associated subtypes of HNC [6], survival rates have improved, as HPV-associated tumors are more sensitive to radiation therapy than HPV-negative tumors and exhibit improved tumor control [7–9]. Moreover, since patients with HPV positive tumors are typically younger and healthier, they have a higher cumulative lifetime risk for ORN development, highlighting the importance of dedicated strategies aimed ORN prevention in modern practice.

ORN is characterized by non-healing bone and mucosal insult following radiation treatment and the condition may present with variable severity [2,10]. Some cases of ORN may clinically heal spontaneously over time (Grade I), while other presentations of ORN may require minor debridement of the injured tissue (Grade II), hyperbaric therapy (Grade III), or major invasive mandible surgery (Grade IV) [11]. Due to the characteristic presence of devitalized bone and reduced blood supply, successful treatment for ORN may be challenging and unpredictable, thus the optimal management for the condition is prevention. Normal Tissue Complication Probability (NTCP) prediction of ORN based on dose-volume parameters can guide radiotherapy mandibular dose constraints in an attempt to prevent the development of ORN in HNC patients [12,13]. NTCP may also be used to guide alternative selections for treatment modalities with less distal beam-path toxicity, such as proton therapy [14]. Furthermore, NTCP prediction may be used to identify patients at medium-high risk of ORN in order to prescribe dedicated follow-up imaging for early detection of ORN and intervention prior to advanced stages [15].

At present, no NTCP model has been developed for ORN, yet previous case-control studies have identified a significant relationship between mandibular dose and the development of ORN [1,10,11,16,17]. Many identified the mandible bone volume receiving 50 Gy (V_50Gy_) as the most important volume parameter (V_xGy_), yet dose parameters (D_X%_, minimum dose to X% of the mandibular volume) were not investigated [1,10,11,16,17]. Moreover, pre-radiotherapy dental extractions have also been identified as a risk factor for ORN development [3,11,18]. Some studies observed a significant association between smoking status and ORN [10,11,18], however, others did not observe this correlation [1,3]. Consequently, in the absence of a formal NTCP model with clinical variables, monolithic non-patient-specific dose constraints are used in general practice. Without a usable NTCP model, the confounding effect of clinical variables on dose-toxicity may be obscured.

To this end, the aim of this study is to develop a multivariable NTCP model for the prediction of development of any grade of ORN following radiotherapy in HNC patients. The model considers both dose-volume parameters and risk factors to provide an optimized pre-treatment ORN risk assessment. Secondary study analysis was aimed to develop an NTCP model for the prediction of advanced (grade IV) ORN.

## Materials and methods

### Patients

Subsequent to Institutional Review Board approval [RCR03-0800], retrospective data collection of patient information for cases with proven squamous cell carcinoma of the head and neck cancer were included, if patients received radiotherapy alone, in combination with surgery, or with chemotherapy, with curative intent, between 2005 and 2015 at MD Anderson Cancer Center. These patients were part of a larger “Big Data Radiotherapy HNC” collection effort that is currently being constructed. Patients with previously documented head and neck irradiation, history of salivary gland cancer and patients with a survival or follow-up time of less than a year were excluded from the study. Generally, the radiotherapy tumor prescription dose range was 68–72 Gy for definitive treatment and 60-66 Gy for post-operative indications. In the study period, for primary tumor and upper neck nodal disease, the vast majority received a split-field technique matching a lower anterior neck field and larynx midline block. Alternatively, “whole-field” IMRT was deployed when tumors were located more inferiorly to avoid under-dosing.

### Data extraction and processing

Planned dose distribution and corresponding planning CT were extracted from various planning systems (Pinnacle, Philips Radiation Oncology Systems; Eclipse, Varian Medical Systems; Raystation, RaySearch Laboratories) to standardized DICOM-RT format. The mandibular bone was subsequently auto-segmented with a previously validated multi-atlas-based auto-segmentation (MABAS) using commercial software ADMIRE (research version 1.1, Elekta AB, Stockholm, Sweden) [19]. Dose-volume histogram (DVH) parameters were extracted with bulk extraction using an in-house-developed software script in MATLAB (version R2014a).

### NTCP endpoints

The primary NTCP endpoint of this study was binary ORN (ORN_I-IV_) development any time point after treatment, in patients with a minimum of 12 months of post-radiotherapy follow-up. The secondary NTCP endpoint was the development of ORN_IV_ (ORN_IV_) at any time point after treatment. The ORN grades are defined as follows [11]: Grade I: minimal bone exposure requiring conservative management; Grade II: bone exposure requiring minor debridement received; Grade III: hyperbaric oxygen needed; Grade IV: major invasive surgery required. ORN cases and grades were identified through querying radiology HNC radiotherapy CT scan reports from the Radiology Information Systems (RIS), together with a thorough manually inspecting the electronic health record.

### Candidate predictors

Candidate mandibular bone DVH variables were: mean, minimum and maximum dose; D_2%_; from D_5%_ – with increments of 5% – to D_95%_; D_97%_; D_98%_; D_99%_; from V_5Gy_ – with increments of 5Gy – to V_70Gy_. The following clinical variables were considered: *age*; *sex* (female vs. male); *tumor subsite* (oral cavity vs. oropharynx vs. hypopharynx/unknown-primary/larynx/nasopharynx: as discrete ordinal 1,2,3); *smoking status* (current vs. former/never); smoking *pack years* (continuous); *PORT* (definitive vs. postoperative radiotherapy); *dental extraction* (no/edentulous vs. dental extractions); and *chemotherapy* (no vs. chemotherapy).

### Statistical modeling

The complete retrospective collected data was randomly divided into a training set and an independent test set with a 70:30 ratio. Univariable logistic regression analysis was performed on the training set to investigate statistically significant DVH and clinical variables (p<0.05). Multivariable NTCP model development was performed with all candidate variables with step-wise forward selection with ranking based on Akaike Information Criterion (AIC) score, while testing per variable selection “step” for significance of p<0.01 with likelihood-ratio test for nested model comparison. The internal validity of the variable selection was estimated by repeating the variable selection 5000 times with a bootstrap procedure (i.e. with replacement), as suggested by the TRIPOD statement [20]. Internal model robustness of variable selection was confirmed if variables were serially selected in the bootstrapped samples. These analyses were performed for the primary (ORN_I-IV_) and secondary endpoint (ORN_IV_) *separately* on the training cohort. Final models were independently validated (i.e. not changing variables and coefficients) using the embargoed test subset. Model performance used area under the ROC (receiver operating characteristic) curve (AUC), Nagelkerke’s R^2^ and the discrimination slope as evaluative criteria. In addition, nested model improvement was determined with AIC score difference (∆) which was considered “significant” when ∆AIC > 2, and ‘strong’ discriminatory/informativeness assertion can be made when ∆AIC > 5. The R-packages Regression Modelling Strategies (version 4.3-1) [21] were implemented for these purposes.

## Results

### Patients

Of the total 1789 HNC patients, 1259 patients were included in this study after screening (Inclusion diagram in Appendix 1), which were randomly split in a training set of 882 patients (70%) and validation set of 377 patients (30%). Median follow-up time for all patients was 57 months (range 12–174). Patient characteristics and demographic are detailed in Table 1. Briefly, the vast majority were oropharyngeal cancer (OPC) patients (66%), followed by oral cavity cancer (15%) and laryngeal cancer patients (13%). The majority were male (83%) treated with IMRT (71%). Demographics were not significantly different between the training and validation set with the exception of sex (p=0.02). From the total cohort, 13.7% (n=173 patients) developed any grade ORN (primary endpoint) and 5% ORN_IV_ (secondary endpoint). Median time to development of ORN was 17 months (range 2–142) post-radiotherapy. The distribution of ORN grades was as follows: grade I (12.7%); grade II (20.8%); grade III (28.9%); and grade IV (n = 37.6%).

**Table 1.** Demographics

|  | Total |  | Training set |  | Validation set |  | p-value |
| --- | --- | --- | --- | --- | --- | --- | --- |
| n (%) | 1259 | (100) | 882 | (70) | 377 | (30) |  |
| **ORN Grade (%)** |  |  |  |  |  |  | 0.681 |
| Any | 173 | (14) | 124 | (14) | 49 | (13) |  |
| G1 | 22 | (2) | 14 | (2) | 8 | (2) | 0.44 |
| G2 | 36 | (3) | 30 | (3) | 6 | (2) |  |
| G3 | 50 | (4) | 36 | (4) | 14 | (4) |  |
| G4 | 65 | (5) | 44 | (5) | 21 | (6) |  |
| **Mean mandible dose (SD)** | 37.74 | (12.51) | 37.45 | (12.86) | 38.41 | (11.67) | 0.217 |
| **Sex (%)** |  |  |  |  |  |  | **0.021** |
| Female | 215 | (17) | 136 | (15) | 79 | (21) |  |
| Male | 1044 | (83) | 746 | (85) | 298 | (79) |  |
| **Age (SD)** | 60.72 | (10.07) | 60.82 | (9.89) | 60.68 | (10.15) | 0.824 |
| **Tumor site (%)** |  |  |  |  |  |  | 0.375 |
| Oral Cavity | 190 | (15) | 54 | (6) | 136 | (36) |  |
| Oropharynx | 826 | (66) | 249 | (28) | 577 | (153) |  |
| Larynx | 159 | (13) | 42 | (5) | 117 | (31) |  |
| Hypopharynx | 24 | (2) | 7 | (1) | 17 | (5) |  |
| Nasopharynx | 22 | (2) | 10 | (1) | 12 | (3) |  |
| Unknown primary | 38 | (3) | 15 | (2) | 23 | (6) |  |
| **T stage (%)** |  |  |  |  |  |  | 0.748 |
| Tx | 10 | (1) | 3 | (0) | 7 | (2) |  |
| T0 | 38 | (3) | 15 | (2) | 23 | (6) |  |
| T1 | 268 | (21) | 76 | (9) | 192 | (51) |  |
| T2 | 416 | (33) | 121 | (14) | 295 | (78) |  |
| T3 | 272 | (22) | 87 | (10) | 185 | (49) |  |
| T4 | 255 | (20) | 75 | (9) | 180 | (48) |  |
| **N stage (%)** |  |  |  |  |  |  | 0.496 |
| N0 | 248 | (20) | 69 | (8) | 179 | (47) |  |
| N1 | 146 | (12) | 51 | (6) | 95 | (25) |  |
| N2 | 834 | (66) | 247 | (28) | 587 | (156) |  |
| N3 | 31 | (2) | 10 | (1) | 21 | (6) |  |
| **p16 HPV positive (%)** |  |  |  |  |  |  | 0.921 |
| positive | 397 | (32) | 281 | (32) | 116 | (31) |  |
| negative | 71 | (6) | 50 | (6) | 21 | (6) |  |
| unknown | 791 | (63) | 551 | (62) | 240 | (64) |  |
| **Technique (%)** |  |  |  |  |  |  | 0.053 |
| 3D-CRT | 123 | (10) | 88 | (10) | 35 | (9) |  |
| IMRT | 891 | (71) | 608 | (69) | 283 | (75) |  |
| VMAT | 224 | (18) | 173 | (20) | 51 | (14) |  |
| IMPT | 21 | (2) | 13 | (1) | 8 | (2) |  |
| **Chemotherapy (%)** |  |  |  |  |  |  | 0.965 |
| No chemotherapy | 233 | (19) | 164 | (19) | 69 | (18) |  |
| Concurrent | 624 | (50) | 436 | (49) | 188 | (50) |  |
| Induction + concurrent | 285 | (23) | 197 | (22) | 88 | (23) |  |
| Induction | 97 | (8) | 70 | (8) | 27 | (7) |  |
| missing | 20 | (2) | 15 | (2) | 5 | (1) |  |
| **Surgery (%)** |  |  |  |  |  |  | 1.000 |
| Definitive | 1043 | (83) | 731 | (83) | 312 | (83) |  |
| Post-operative | 216 | (17) | 151 | (17) | 65 | (17) |  |
| **Dental Status (%)** |  |  |  |  |  |  | 0.212 |
| no extraction | 707 | (56) | 506 | (57) | 201 | (53) |  |
| edentulous | 210 | (17) | 137 | (16) | 73 | (19) |  |
| dental extraction | 342 | (27) | 239 | (27) | 103 | (27) |  |
| **Smoking status (%)** |  |  |  |  |  |  | 0.49 |
| Current | 180 | (14) | 121 | (14) | 59 | (16) |  |
| Former | 607 | (48) | 434 | (49) | 173 | (46) |  |
| Never | 472 | (37) | 327 | (37) | 145 | (38) |  |
| **Pack years (SD)** | 20.39 | (28.62) | 21.84 | (28.89) | 19.77 | (28.50) | 0.238 |

Abbreviations: SD: standard deviation; HPV: Human Papilloma Virus

### Univariable analyses

All DVH parameters were significantly associated with the development of ORN (any grade) in univariable analyses. The parameters ranging between D_2%_-D_98%_ and V_15Gy_-V_70Gy_ were highly significant (p<0.0001) (Appendix 2). The D_30%_ and V_50Gy_ showed the best classification performance with AUCs of 0.76 (95% Confidence Interval(95%CI): [0.72-0.80]). Notably, DVH parameters ranging from D_15%_ to D_55%_, V_40Gy_ to V_60Gy_ and mean mandible dose performed similarly (AUC: [0.74-0.76]). Figure 1 depicts the dose (D_x%_) and volume (V_xGy_) distinction of patients that do and do not develop ORN and the DVH parameter significance level. For example, Figure 1 showed that patients that did not develop ORN received an average D_30%_ of 46±16Gy, while this was 57±9Gy for those who did develop ORN. Additionally, dental extraction (Odds ratio (OR)=1.67 (1.35-2.06); p<0.0001), *PORT* (OR=1.68 (1.07-2.65); p=0.0253) and chemotherapy (OR=1.85 (1.06-3.21); p=0.0293), were significantly associated with the development of ORN (Table 2). Specifically for tumor site, compared to *oral cavity* (i.e. as reference), ORs were 0.58 [0.37-0.92] (p=0.021) for *OPC* and 0.08 [0.03-0.23] (p<0.0001) for *others*. Both *smoking status* and *pack years* did not show a significant relationship with ORN development.

**Table 2.**
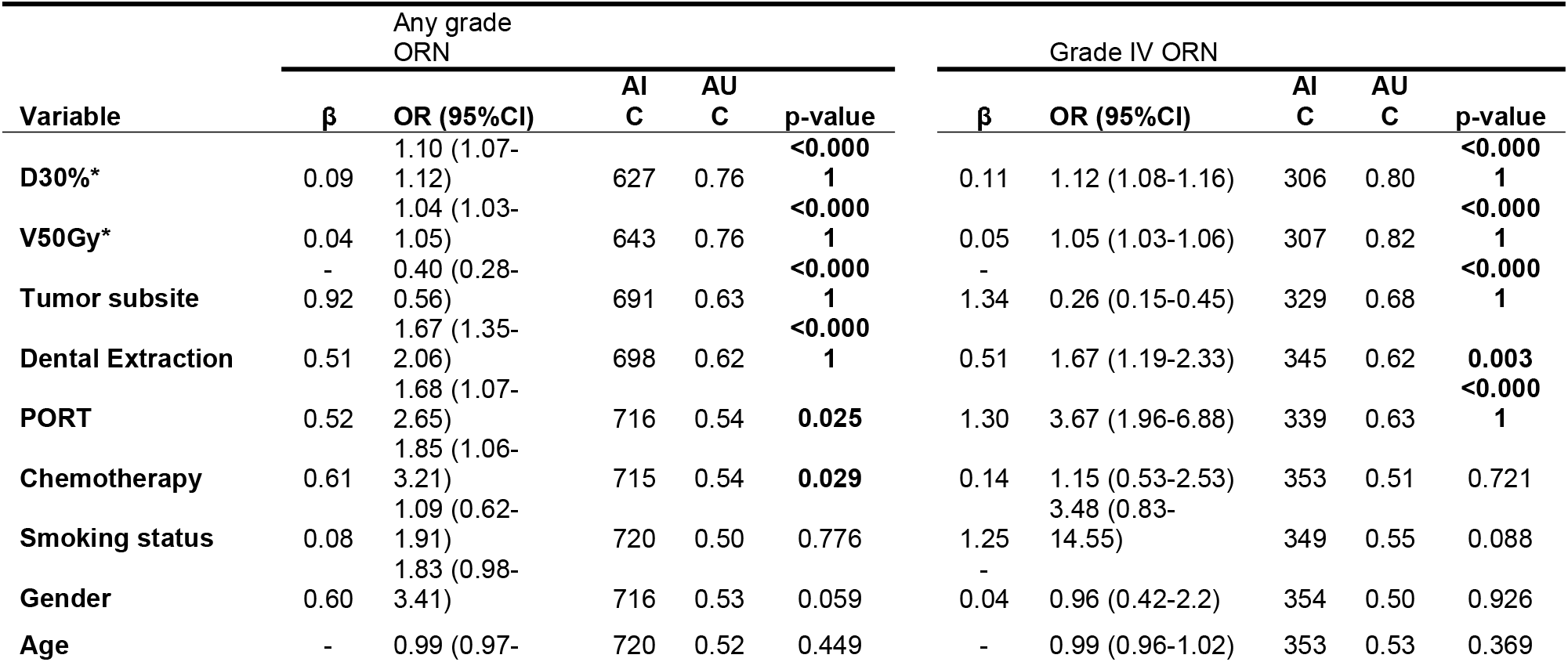
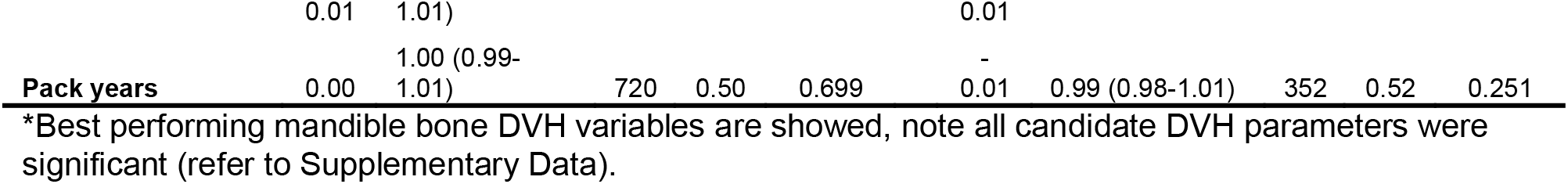
Univariable results of best performing dose volume parameters, all clinical variables

**Figure 1.**
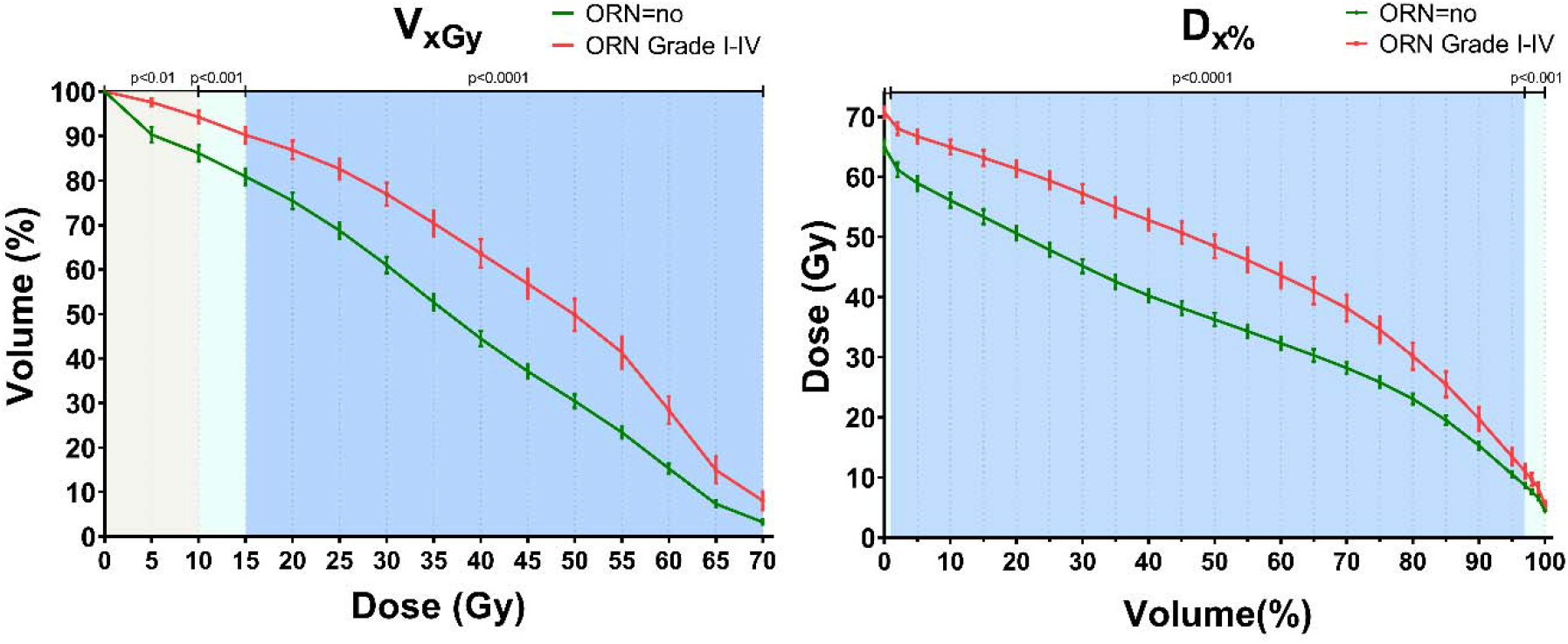
Average DVH for patient that develop ORN (red) versus those who do not (green) for volume (V_xGy_) (left) and dose (D_x%_) parameters (right). Color shading indicates the univariable significance of parameters, indicating that D_2%_-D_98%_ and V_15Gy_-V_70Gy_ were significant with a p<0.0001.

### Multivariable NTCP model development and validation

AIC-ranked forward selection in the training set step-wise identified D_30%_ first (p<0.0001) followed by *pre-RT dental extraction* (likelihood-ratio test; p=0.005) with a “significance” ∆AIC of 5.96. Bootstrapped forward variable selection in the training cohort also showed that D_30%_ was the most frequently selected first variable (50% of the bootstrapped samples; note, D_25%_ in 23%) and the clinical variable *dental extraction* as the second variable (47%; Appendix 3). The positive regression coefficients reveal that higher D_30%_ (OR=1.10 [1.07-1.12]) and the *dental extraction* (OR=1.67 [1.35-2.06]) are associated with higher risk of developing ORN (Table 3). The model performance was good with an AUC of 0.78 [0.74-0.82] and R^2^ of 0.20 (Table 4). Validation of the performance of the NTCP model with D_30_ and *dental extraction* was tested on the independent test set (n=377) was also good (AUC_validation_=0.75; R^2^=0.17).

**Table 3.** Model parameter for any grade (Grade I-IV) and Grade IV ORN NTCP models.

| Variables | Grade I-IV ORN |  |  | Grade IV ORN |  |  |  |  |  |
| --- | --- | --- | --- | --- | --- | --- | --- | --- | --- |
| | $\beta$ | OR | p-value | $\beta$ | OR | p-value | $\beta$ | OR | p-value |
| **intercept** | -6.85 |  |  | -9.16 |  |  | -12.27 |  |  |
| **D30** | 0.09 | 1.1 (1.07-1.12) | <0.0001 | 0.11 | 1.12 (1.07-1.16) | <0.0001 | 0.12 | 1.13 (1.08-1.17) | <0.0001 |
| **Dental extractions** | 0.66 | 1.93 (1.28-2.92) | 0.002 | 0.62 | 1.85 (0.98-3.49) | 0.057 |  |  |  |
| **Smoking status** |  |  |  |  |  |  | 1.51 | 4.54 (1.05-19.68) | 0.043 |

For the secondary NTCP endpoint ORN_IV_ (i.e. needing major surgical intervention), forward selection selected the dose variable D_30%_. Disregarding dose variables (V_70/65Gy_) that flipped to negative coefficient in multivariable analyses (i.e. suggesting over/incorrect-fitting), smoking status was the next most-associated variable, but did not meet our pre-specified significance level (likelihood-ratio test; p=0.013), nor did dental extraction (p=0.06). Bootstrapped variable selection selected V_55Gy_ (32%) over D_25%_ (20%), D_30%_ (18%), D_40%_ (14%) (Appendix 3), together with the clinical variables smoking status (29%) and/or dental extraction (21%) in multiple “runs”. In training, ORN_IV_ model performance was nearly identical for NTCP models with D_30%_ or V_55Gy_ alone, or combined with *smoking status* or *dental extraction* (AUC range: 0.80-0.82; R^2^ range: 0.16-0.18). However, external validation showed that the model with D_30%_ and *dental extraction* (AUC_validation_=0.82; R^2^_validation_=0.21) performed marginally better than models with V_55Gy_ or *smoking status* (refer to Appendix 4). Performance improved with a model D_30%_ and *dental extraction* compared to the model with D_30%_ alone; even though this improvement is limited, for consistency with ORN_I-IV_, we selected the same two variables in the final ORN_IV_ NTCP model (note: coefficients deviate).

Final NTCP models that were developed in the training cohort (model coefficients in Table 3) and validated in the unseen/embargoed test cohort are plotted in Figure 2. Binned actual observed ORN proportions, represented by points with error bars, correspond with the NTCP models. The horizontal gray lines in Figure 2 indicate the 5% ORN threshold risks.

**Figure 2.**
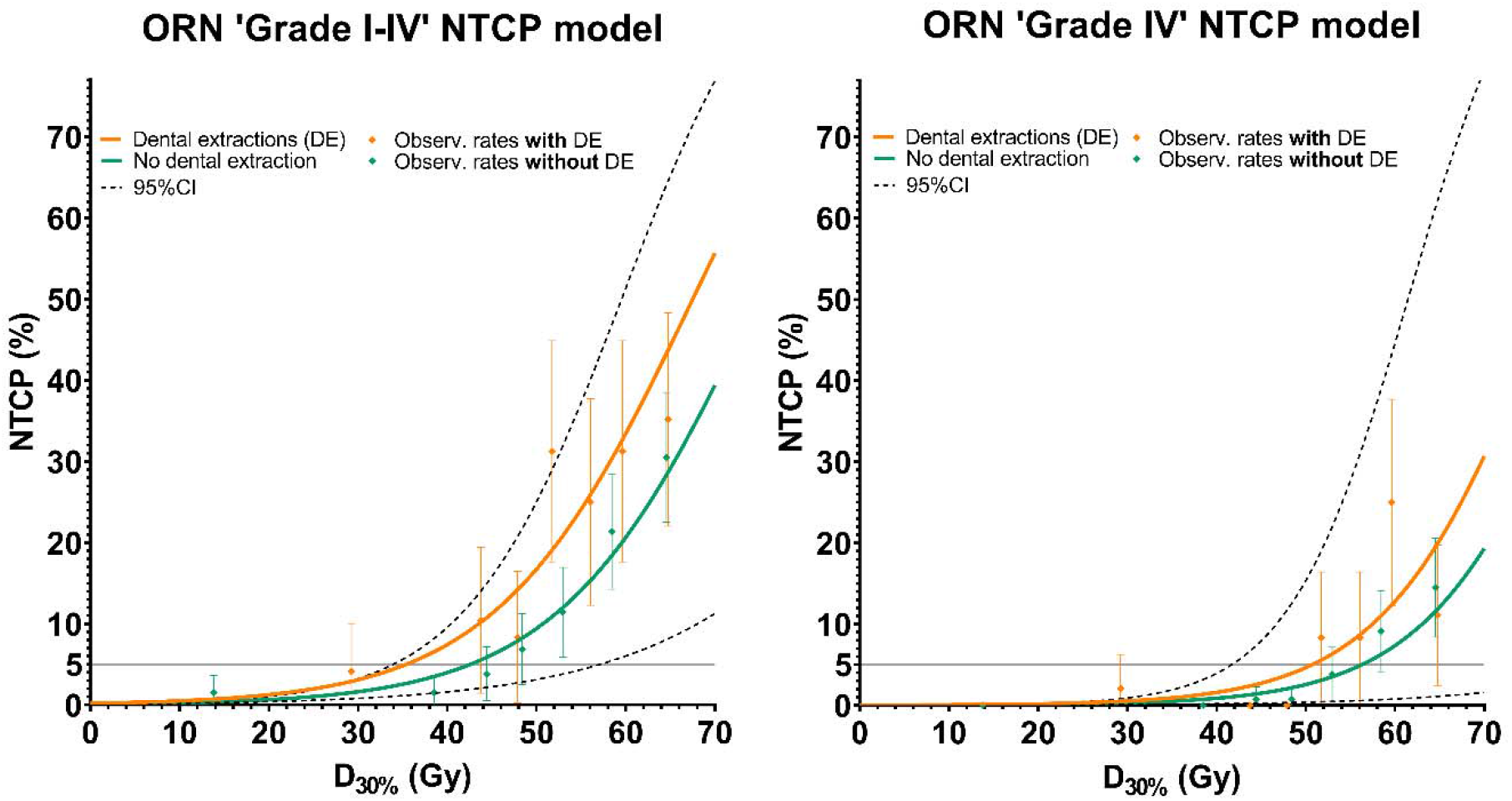
Final ORN NTCP models. NTCP curves are plotted against D_30%_ split into patient with pre-treatment dental extraction (orange lines) and those without (green lines) for NTCP models for ORN any grade (left plot) and Grade IV (right). Point with error bars of the actual observed ORN rates by binning all patients sorted by dose in X percentiles, points are positioned at the average dose per bin.

### Sub-cohort analyses

The final NTCP model (Table 3) performed similarly for OPC patients only (n=826; AUC_OPC- cohort_=0.76), for larynx/hypo/nasopharynx/unknown-primary cancer patients (i.e. others; n=243; AUC_Other-cohort_=0.76), and combined cohorts (i.e. OPC+others patients; n=1069; AUC_OPC+other-cohort_=0.79). In contrast, performance in the oral cavity cancer patients was poor (n=190; AUC_Oral cavity_=0.59). A similar trend was seen for ORN_IV_ (AUC_OPC-cohort_=0.80, AUC_OPC+ther-cohort_=0.84, AUC_Oral cavity_=0.57), except that in the ‘others cohort’ no ORN_IV_ was present. Refer to Appendix 5 for sub-analyses test results per tumor site, and for definitive and PORT patients.

## Discussion

While ORN rates are relatively low (~5-15%), the consequences for patients experiencing ORN are highly disabling, with a substantial impact on the healthcare utilization and quality of life [5]. Once ORN develops, treatment is complicated by the lack of regenerative bone and tissue cells needed for healing and repair. Advanced stage ORN requires extensive surgery associated with significant peri-operative morbidity [22]. Given the potential severity of ORN and limitations in treatment once developed, improved pre-treatment risk assessment tools aimed at identifying high-risk patients and guiding strategies for prevention and early intervention of ORN represent an important unmet need.

Due to the low relative prevalence of ORN, large-scale radiation dose and matching toxicity data is needed. At present, previous studies assessing radiation dose to the mandible and development of ORN have at best 200-600 patients [1,10,11,16,17]. In response to the unmet need for prediction models for the development of ORN and ORN severity validated across data from a sufficient patient cohort, this study developed NTCP models for the prediction of ORN of *any grade* and *Grade IV* in a large cohort of 1259 HNC patients treated with definitive or post-operative (chemo-) radiotherapy.

The association between ORN development and mandible radiation dose was clearly observed with the univariable significance of all DVH parameters (Figure 1). The final NTCP models were based on *D*_*30%*_, which represents the dose to 30% of the mandibular volume, and *pre-treatment dental extraction*. This NTCP model had good performance in both the training and validation cohort for ORN_I-IV_ (AUC_train/validation_=0.78/0.75) and ORN_IV_ (AUC_train/validation_=0.81/0.82). These models are clinically useful tools to determine appropriate dose constraints for the mandibular bone when feasible (i.e. when tumor coverage is not compromised) [14]. Additionally, they identify patients at high risk for ORN development, who require more intensive clinical surveillance programs with dedicated imaging follow-up [23] and/or earlier intervention, whether conservative or surgical, to prevent ORN progression.

Our results demonstrate that mandible dose constraints can be distilled from these NTCP models in order to optimize patients’ IMRT plans. For example, our models suggest that mandibular D_30%_ of patients without pre-treatment dental extraction should be kept below 42Gy to achieve <5% risk of ORN development, while a D_30%_<35Gy is required for patients with dental extractions to achieve the same level of risk (Figure 2). Alternately, for a more conservative risk threshold of 1%, D_30%_ should be <25Gy (without dental extractions) and <17Gy (dental extractions). In respect to ORN_IV_ only, maintaining D_30%_<56Gy without pre-RT dental extractions or D_30%_<50Gy with pre-RT dental extractions may be sufficient to achieve <5% risk of ORN_IV_ development.

Our findings of significant association between ORN and several DVH parameters as well as with pre-dental status match with the results of several recent publications [1,10,11,16,17,24]. For instance, a recent publication from a Danish group showed that several DVH parameters in the intermediate and high dose range including D_mean_ were associated with ORN in a cohort of HNC patients with 56 ORN cases and 112 controls [24]. Another study from the Princess Margaret Cancer Centre, reported that V_50_ and V_60_ were significantly higher in 71 ORN patients compared to 142 patients with no ORN [10]. In addition, another group previously reported that maximum radiation dose to the mandible as a single dose constraint was a poor correlate of ORN in OPC patients, and that mandibular volumes receiving 44 Gy (V_44Gy_), and 58 Gy (V_58Gy_) were comparatively more discriminatory of ORN versus non-ORN patients [1]. However, these studies were case-control studies, were based on limited number of patients, and did not investigate dose parameters (D_x%_), recommend risk-based DVH constraints, nor design a multivariable NTCP model.

Our multivariable NTCP models showed that a combination of mandibular dosimetric parameters (D_30%_) with the pre-RT dental extraction status achieve the best performing model for ORN risk prediction. A study by the Memorial Sloan Kettering Cancer Center group showed that, in addition to mandibular radiation doses, the presence of mild-severe periodontal bone loss was associated with increased ORN risk. However, in this study, pre-RT radiographs were only available for 18 ORN patients that were matched with 36 controls [3]. In concurrence with our study results, several recent studies have demonstrated that pre-radiotherapy dental, rather than post-radiotherapy, extraction was a significant risk factor for ORN development [10,18,25–27].

Whether pre-radiotherapy dental extraction is a direct incipient insult preceding ORN development or merely a surrogate for poor dentition remains unclear. While all patients receive pre-therapy dental oncology assessment, we do not routinely deploy asymptomatic dental surveillance, referring these cases to their community dentists. Consequently, our data set lacked significant prospective post-radiation dental assessment variables, surveillance of radiation caries, and post-therapy dental extraction outside our facility extraction was not captured. Consequently, there remains a significant need to undertake prospective assessment of orodental health with developed instruments (e.g. formal sialometry, radiation caries monitoring with DMFS160 [28] and patient-reported outcomes) to determine whether the observed association of ORN with pre-therapy dental extractions can be related to one or more mechanisms.

In contrast to previous studies [10,11,18], *smoking status* was not found to be significantly associated with all grades of ORN_I-IV_ in the current study, but *smoking status* was frequently identified on variable selection with higher ORN grade (i.e. grade IV). Notably, our validation showed reduced performance of models with *smoking status* included compared to that in the training cohort – in contrast to the model with *dental extraction*. Other groups have shown similar ambiguity as to the role of tobacco on ORN, with other publications also showing no association between smoking status and ORN [1,3]. These contradictory findings may be due to inter-cohort variables inherent in different studies’ population. A second possible explanation is that smoking continuation during and after treatment may be of more influence for the development of ORN compared with patients who elect to stop smoking before treatment, as in our dataset, which had limited active smokers. More research is needed to investigate the discordance between our findings and other group reports [10,11,18].

Sub-analyses showed that in the NTCP model performance was poor when tested in the oral cavity cancer patients only (AUC_Oral-cavity_=0.59), especially relative to the performance in the non-oral cavity cancer patients (AUC_OPC+Other-cohort_=0.79). For the oral cavity cancer patients, both the mandible dose (D_mean_= 46.5±6.5 Gy) and ORN prevalence (23%) was higher compared to the rest of the cohort (36.2±12.7 Gy; prevalence:12%). While relatively small oral cavity cancer patient sample size (n=190) could explain the limited significance of the dose variables (i.e. only D_30%_, D_35%,_ and D_40%_), the model poor performance of the NTCP models suggests that there is an effect in these patients not captured in the present dataset. One consideration is that oral cavity cancer patients typically received PORT (89%), while patients with other tumor sites were generally treated with primary RT (94%). Across tumor locations, NTCP model performance was better in patients treated with definitive radiotherapy group (AUC=0.78) than the PORT group (AUC=0.65) (Appendix 5). While *PORT* was significant in univariable analysis, it did not perform well in the multivariable analyses. Further research with specific focus on role of pre-RT surgical intervention and/or other oral cavity-specific factors is needed to better explain ORN development in oral cavity cancer patients. Additionally, while oral cavity cancer patients generally receive radiation to greater volumes of the mandible, the gradients across the mandible were more homogenous. The current NTCP approach treats DVH dose-volume “bins” as discrete independent constructs which may obfuscate discriminatory signal in organs with more homogenous cohort dose distributions and suggests further investigation with alternative normal tissue injury approaches are warranted.

Though this study is based on an extensive retrospective cohort of HNC patients treated between 2005 and 2015 at MD Anderson Cancer Center, limitations include that this sample represents a fraction all patients in the study time frame, an estimated 25%. Nevertheless, we are convinced that the included patient cohort is likely a fair representation of our institutional HNC population. Moreover, we considered ORN development as a binary variable, leading (as in most NTCP studies) to potential limitations with regard to right-censored event prediction. For simplicity, we used conventional NTCP model approaches, but future efforts at dynamic time-incorporating risk models (e.g. partially observed Markov decision processes) are ongoing.

Despite these limitations, to our knowledge this study represents the largest extant ORN survey of dose-response data, and the first published ORN NTCP model. To ensure FAIR data [29] and allow external validation, an anonymized version of the dataset, including DVH and clinical variables with ORN grades have been deposited at [doi:10.6084/m9.figshare.13568207]. Our hope is that this can afford others the opportunity to validate our approach, generate institutional-specific models, and engender further cross-platform research for ORN toxicity modeling, and multi-institutional dose constraints.

## Conclusion

The developed NTCP models performed well in predicting ORN_I-IV_ (primary NTCP endpoint) and ORN_IV_ (secondary NTCP endpoint) in both the HNC patient training and independent test cohort. NTCP models were based on mandible bone D_30%_ and pre-treatment *dental extraction*. Our results show a distinct association between planned mandible bone radiation dose and ORN development, and suggest that less than 30% of the mandible should receive a dose of 35Gy or more for an ORN_I-IV_ risk lower than 5%. These NTCP models may be utilized to improve prevention of ORN as well as guide ORN surveillance/management strategies by identifying and stratifying patients at risk of ORN.

## Data Availability

To ensure FAIR data and allow external validation, an anonymized version of the dataset, including DVH and clinical variables with ORN grades have been deposited at [doi:10.6084/m9.figshare.13568207].

doi:10.6084/m9.figshare.13568207

## Notes

### Competing Interest Statement

The authors have declared no competing interest.

### Funding Statement

Dr. van Dijk, received/receives funding and salary support from the Dutch organization NWO ZonMw during the period of study execution via the Rubicon Individual career development grant.

Drs. Mohamed and Abusaif are funded for this work by The University of Texas MD Anderson Cancer Center-Oropharynx Cancer Program generously supported by Mr. and Mrs. Charles W. Stiefel.

Dr. Hutcheson, Mohamed, Lai, and Fuller received/receives funding and salary support related to this project during the period of study execution from: the NIH National Cancer Institute (NCI) Early Phase Clinical Trials in Imaging and Image-Guided Interventions Program (R01CA218148).

Dr. Lai, Mohamed, and Fuller received/receives funding and salary support related to this project during the period of study execution from: the National Institutes of Health (NIH) National Institute for Dental and Craniofacial Research (NIDCR) Establishing Outcome Measures Award (R01DE025248/R56DE025248).

Dr. Fuller received funding unrelated to this project during the period of study execution from NIH/NCI Cancer Center (P30CA016672, P50 CA097007, and R01CA2148250); from NIH/NIBIB (R25EB025787-01); from NIH/NSF (NSF1557679); NSF-CMMI grant (NSF1933369); and the Sabin Family Foundation.

### Author Declarations

MD Anderson Cancer Centers Institutional Review Board gave approval for the retrospective collection of imaging, dose and clinical information of head and neck cancer patients at MD Anderson Cancer Center [RCR03-0800]

## References

[1] Mohamed ASR, Hobbs BP, Hutcheson KA, Murri MS, Garg N, Song J, et al. Dose-volume correlates of mandibular osteoradionecrosis in Oropharynx cancer patients receiving intensity-modulated radiotherapy: Results from a case-matched comparison. Radiother Oncol 2017;124:232–9.

[2] Chronopoulos A, Zarra T, Ehrenfeld M, Otto S. Osteoradionecrosis of the jaws: definition, epidemiology, staging and clinical and radiological findings. A concise review. Int Dent J 2018;68:22–30.

[3] Owosho AA, Tsai CJ, Lee RS, Freymiller H, Kadempour A, Varthis S, et al. The prevalence and risk factors associated with osteoradionecrosis of the jaw in oral and oropharyngeal cancer patients treated with intensity-modulated radiation therapy (IMRT): The Memorial Sloan Kettering Cancer Center experience. Oral Oncol 2017;64:44–51.

[4] Beadle BM, Liao K-P, Chambers MS, Elting LS, Buchholz TA, Kian Ang K, et al. Evaluating the impact of patient, tumor, and treatment characteristics on the development of jaw complications in patients treated for oral cancers: A SEER-Medicare analysis. Head Neck 2013;35:1599–605.

[5] Wong ATT, Lai SY, Gunn GB, Beadle BM, Fuller CD, Barrow MP, et al. Symptom burden and dysphagia associated with osteoradionecrosis in long-term oropharynx cancer survivors: A cohort analysis. Oral Oncol 2017;66:75–80.

[6] Siegel RL, Miller KD, Jemal A. Cancer statistics, 2020. CA Cancer J Clin 2020;70:7–30.

[7] Licitra L, Perrone F, Bossi P, Suardi S, Mariani L, Artusi R, et al. High-risk human papillomavirus affects prognosis in patients with surgically treated oropharyngeal squamous cell carcinoma. J Clin Oncol 2006;24:5630–6.

[8] Sedaghat AR, Zhang Z, Begum S, Palermo R, Best S, Ulmer KM, et al. Prognostic significance of human papillomavirus in oropharyngeal squamous cell carcinomas. Laryngoscope 2009;119:1542–9.

[9] Ang KK, Harris J, Wheeler R, Weber R, Rosenthal DI, Nguyen-Tân PF, et al. Human Papillomavirus and Survival of Patients with Oropharyngeal Cancer. N Engl J Med 2010;363:24–35.

[10] Caparrotti F, Huang SH, Lu L, Bratman S V., Ringash J, Bayley A, et al. Osteoradionecrosis of the mandible in patients with oropharyngeal carcinoma treated with intensity-modulated radiotherapy. Cancer 2017;123:3691–700.

[11] Tsai CJ, Hofstede TM, Sturgis EM, Garden AS, Lindberg ME, Wei Q, et al. Osteoradionecrosis and radiation dose to the mandible in patients with oropharyngeal cancer. Int J Radiat Oncol Biol Phys 2013;85:415–20.

[12] Kierkels RGJ, Korevaar EW, Steenbakkers RJHM, Janssen T, Van’T Veld AA, Langendijk JA, et al. Direct use of multivariable normal tissue complication probability models in treatment plan optimisation for individualised head and neck cancer radiotherapy produces clinically acceptable treatment plans. Radiother Oncol 2014;112:430–6.

[13] Witte MG, Van Der Geer J, Schneider C, Lebesque J V., Alber M, Van Herk M. IMRT optimization including random and systematic geometric errors based on the expectation of TCP and NTCP. Med Phys 2007;34:3544–55.

[14] Langendijk JA, Lambin P, De Ruysscher D, Widder J, Bos M, Verheij M. Selection of patients for radiotherapy with protons aiming at reduction of side effects: the model-based approach. Radiother Oncol 2013;107:267–73.

[15] Sandulache VC, Hobbs BP, Mohamed ASR, Frank SJ, Song J, Ding Y, et al. Dynamic contrast-enhanced MRI detects acute radiotherapy-induced alterations in mandibular microvasculature: prospective assessment of imaging biomarkers of normal tissue injury. Sci Rep 2016;6:29864.

[16] Zhang W, Zhang X, Yang P, Blanchard P, Garden AS, Gunn B, et al. Intensity-modulated proton therapy and osteoradionecrosis in oropharyngeal cancer. Radiother Oncol 2017;123:401–5.

[17] Brodin NP, Tomé WA. Revisiting the dose constraints for head and neck OARs in the current era of IMRT. Oral Oncol 2018;86:8–18.

[18] Moon DH, Moon SH, Wang K, Weissler MC, Hackman TG, Zanation AM, et al. Incidence of, and risk factors for, mandibular osteoradionecrosis in patients with oral cavity and oropharynx cancers. Oral Oncol 2017;72:98–103.

[19] Mohamed ASR, Ruangskul M-N, Awan MJ, Baron CA, Kalpathy-Cramer J, Castillo R, et al. Quality Assurance Assessment of Diagnostic and Radiation Therapy–Simulation CT Image Registration for Head and Neck Radiation Therapy: Anatomic Region of Interest– based Comparison of Rigid and Deformable Algorithms. Radiology 2015;274:752–63.

[20] Moons KGM, Altman DG, Reitsma JB, Ioannidis JPA, Macaskill P, Steyerberg EW, et al. Transparent Reporting of a multivariable prediction model for Individual Prognosis Or Diagnosis (TRIPOD): Explanation and Elaboration. Ann Intern Med 2015;162:W1.

[21] R Development Core Team. R: A Language and Environment for Statistical Computing. Vienna, Austria: the R Foundation for Statistical Computing. 2011:Available online at http://www.R-project.org/.

[22] Lambade PN, Lambade D, Goel M. Osteoradionecrosis of the mandible: A review. Oral Maxillofac Surg 2013;17:243–9.

[23] Head J, Cooperative NRD. Dynamic contrast-enhanced MRI detects acute radiotherapy-induced alterations in mandibular microvasculature: prospective assessment of imaging biomarkers of normal tissue injury. Sci Rep 2016;6:29864.

[24] Aarup-Kristensen S, Hansen CR, Forner L, Brink C, Eriksen JG, Johansen J. Osteoradionecrosis of the mandible after radiotherapy for head and neck cancer: risk factors and dose-volume correlations. Acta Oncol (Madr) 2019;58:1373–7.

[25] Beech NM, Porceddu S, Batstone MD. Radiotherapy-associated dental extractions and osteoradionecrosis. Head Neck 2017;39:128–32.

[26] Sathasivam HP, Davies GR, Boyd NM. Predictive factors for osteoradionecrosis of the jaws: A retrospective study. Head Neck 2018;40:46–54.

[27] Kojima Y, Yanamoto S, Umeda M, Kawashita Y, Saito I, Hasegawa T, et al. Relationship between dental status and development of osteoradionecrosis of the jaw: a multicenter retrospective study. Oral Surg Oral Med Oral Pathol Oral Radiol 2017;124:139–45.

[28] Watson E, Eason B, Kreher M, Glogauer M. The DMFS160: A new index for measuring post-radiation caries. Oral Oncol 2020;108:104823.

[29] Goble C, Cohen-Boulakia S, Soiland-Reyes S, Garijo D, Gil Y, Crusoe MR. FAIR Computational Workflows. Data Intell 2019;23:0–2.

