## Supplementary material for "Normal Tissue Complication Probability (NTCP) prediction model for osteoradionecrosis of the mandible in head and neck cancer patients following radiotherapy: Large-scale observational cohort": Suplementary Data

**Appendix 1**

**Figure.** Patient inclusion diagram

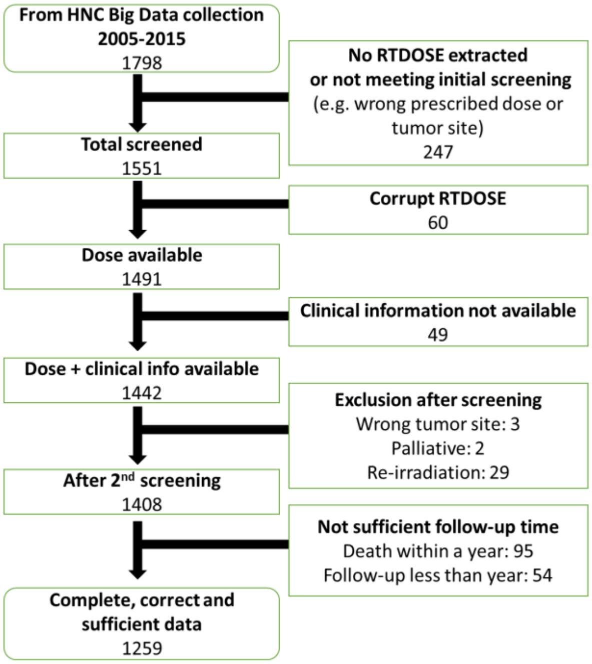

### Appendix 2

**Table.** Full Univariable analysis table for ORN any grade (I-IV) and Grade IV

| | ORN Grade I-IV | | | | | ORN Grade IV | | | | | |
|---|---|---|---|---|---|---|---|---|---|---|---|
| **Variable** | **β** | **OR (95%CI)** | **AIC** | **AUC** | **p-value** | **β** | **OR (95%CI)** | **AIC** | **BIC** | **AUC** | **p-value** |
| **D30** | 0.09 | 1.1 (1.07-1.12) | 627 | 0.76 | **<0.0001** | 0.11 | 1.12 (1.08-1.16) | 306 | 316 | 0.80 | **<0.0001** |
| **D25** | 0.10 | 1.11 (1.08-1.13) | 629 | 0.76 | **<0.0001** | 0.12 | 1.13 (1.08-1.18) | 308 | 317 | 0.80 | **<0.0001** |
| **D35** | 0.09 | 1.09 (1.07-1.11) | 629 | 0.76 | **<0.0001** | 0.10 | 1.11 (1.07-1.15) | 308 | 317 | 0.79 | **<0.0001** |
| **D40** | 0.08 | 1.08 (1.06-1.1) | 631 | 0.76 | **<0.0001** | 0.09 | 1.1 (1.06-1.14) | 308 | 317 | 0.79 | **<0.0001** |
| **D45** | 0.07 | 1.08 (1.06-1.1) | 634 | 0.75 | **<0.0001** | 0.09 | 1.09 (1.06-1.13) | 309 | 318 | 0.79 | **<0.0001** |
| **D20** | 0.10 | 1.11 (1.08-1.14) | 638 | 0.75 | **<0.0001** | 0.12 | 1.13 (1.08-1.19) | 313 | 322 | 0.78 | **<0.0001** |
| **D50** | 0.07 | 1.07 (1.05-1.09) | 641 | 0.74 | **<0.0001** | 0.08 | 1.08 (1.05-1.11) | 313 | 323 | 0.77 | **<0.0001** |
| **mean** | 0.10 | 1.1 (1.07-1.13) | 642 | 0.74 | **<0.0001** | 0.10 | 1.1 (1.06-1.15) | 320 | 330 | 0.75 | **<0.0001** |
| **V45** | 0.04 | 1.04 (1.03-1.05) | 642 | 0.75 | **<0.0001** | 0.05 | 1.05 (1.03-1.06) | 311 | 321 | 0.79 | **<0.0001** |
| **V50** | 0.04 | 1.04 (1.03-1.05) | 643 | 0.76 | **<0.0001** | 0.04 | 1.05 (1.03-1.06) | 309 | 318 | 0.81 | **<0.0001** |
| **V55** | 0.04 | 1.04 (1.03-1.05) | 644 | 0.76 | **<0.0001** | 0.05 | 1.05 (1.03-1.06) | 307 | 316 | 0.82 | **<0.0001** |
| **V40** | 0.04 | 1.04 (1.03-1.05) | 645 | 0.74 | **<0.0001** | 0.05 | 1.05 (1.03-1.07) | 315 | 325 | 0.77 | **<0.0001** |
| **D55** | 0.06 | 1.07 (1.05-1.08) | 646 | 0.74 | **<0.0001** | 0.07 | 1.07 (1.05-1.1) | 318 | 327 | 0.76 | **<0.0001** |
| **D15** | 0.10 | 1.11 (1.07-1.14) | 649 | 0.74 | **<0.0001** | 0.12 | 1.13 (1.07-1.19) | 320 | 329 | 0.75 | **<0.0001** |
| **V35** | 0.04 | 1.04 (1.03-1.05) | 652 | 0.72 | **<0.0001** | 0.05 | 1.05 (1.03-1.07) | 321 | 331 | 0.74 | **<0.0001** |
| **D60** | 0.06 | 1.06 (1.05-1.08) | 653 | 0.73 | **<0.0001** | 0.06 | 1.07 (1.04-1.09) | 323 | 332 | 0.74 | **<0.0001** |
| **D65** | 0.06 | 1.06 (1.04-1.08) | 659 | 0.72 | **<0.0001** | 0.06 | 1.06 (1.04-1.09) | 327 | 337 | 0.73 | **<0.0001** |
| **D10** | 0.10 | 1.1 (1.07-1.14) | 661 | 0.71 | **<0.0001** | 0.11 | 1.11 (1.05-1.17) | 329 | 339 | 0.71 | **0.0001** |
| **V30** | 0.04 | 1.04 (1.03-1.06) | 661 | 0.70 | **<0.0001** | 0.05 | 1.05 (1.02-1.07) | 329 | 339 | 0.71 | **<0.0001** |
| **V60** | 0.04 | 1.04 (1.03-1.05) | 661 | 0.74 | **<0.0001** | 0.04 | 1.04 (1.03-1.06) | 325 | 334 | 0.80 | **<0.0001** |
| **D70** | 0.05 | 1.06 (1.04-1.07) | 665 | 0.70 | **<0.0001** | 0.05 | 1.06 (1.03-1.08) | 331 | 340 | 0.71 | **<0.0001** |
| **V25** | 0.04 | 1.04 (1.03-1.06) | 673 | 0.68 | **<0.0001** | 0.04 | 1.04 (1.02-1.07) | 337 | 347 | 0.67 | **0.002** |
| **D5** | 0.09 | 1.09 (1.05-1.13) | 675 | 0.68 | **<0.0001** | 0.08 | 1.08 (1.03-1.15) | 338 | 348 | 0.65 | **0.004** |
| **D75** | 0.05 | 1.05 (1.04-1.07) | 675 | 0.69 | **<0.0001** | 0.05 | 1.05 (1.02-1.07) | 338 | 347 | 0.68 | **<0.0001** |
| **D80** | 0.05 | 1.05 (1.03-1.06) | 686 | 0.66 | **<0.0001** | 0.04 | 1.04 (1.01-1.06) | 344 | 353 | 0.62 | **0.002** |
| **D2** | 0.07 | 1.07 (1.04-1.11) | 687 | 0.65 | **<0.0001** | 0.06 | 1.06 (1.01-1.12) | 344 | 353 | 0.61 | **0.023** |
| **V20** | 0.04 | 1.04 (1.02-1.05) | 687 | 0.65 | **<0.0001** | 0.03 | 1.03 (1-1.05) | 345 | 355 | 0.61 | **0.021** |
| **V65** | 0.04 | 1.04 (1.03-1.05) | 687 | 0.64 | **<0.0001** | 0.03 | 1.03 (1.01-1.04) | 347 | 357 | 0.58 | **0.005** |
| **D85** | 0.05 | 1.05 (1.03-1.07) | 690 | 0.64 | **<0.0001** | 0.04 | 1.04 (1.01-1.07) | 345 | 354 | 0.61 | **0.003** |
| **type** | -0.92 | 0.4 (0.28-0.56) | 691 | 0.63 | **<0.0001** | -1.34 | 0.26 (0.15-0.45) | 329 | 339 | 0.68 | **<0.0001** |
| **V70** | 0.05 | 1.06 (1.03-1.08) | 691 | 0.62 | **<0.0001** | 0.03 | 1.03 (1-1.06) | 350 | 359 | 0.58 | **0.026** |
| **maxGy** | 0.00 | 1 (1-1) | 696 | 0.62 | **0.0005** | 0.00 | 1 (1-1) | 348 | 358 | 0.57 | **0.076** |
| **V15** | 0.03 | 1.03 (1.01-1.05) | 697 | 0.62 | **0.0002** | 0.02 | 1.02 (1-1.04) | 349 | 359 | 0.56 | **0.075** |
| **D90** | 0.05 | 1.05 (1.03-1.07) | 698 | 0.63 | **<0.0001** | 0.04 | 1.04 (1.01-1.07) | 348 | 358 | 0.60 | **0.016** |
| **V10** | 0.03 | 1.03 (1.01-1.05) | 701 | 0.59 | **0.002** | 0.02 | 1.02 (1-1.04) | 350 | 359 | 0.54 | **0.109** |
| **Dental extraction** | 0.87 | 2.39 (1.61-3.54) | 702 | 0.60 | **<0.0001** | 0.86 | 2.36 (1.28-4.35) | 346 | 356 | 0.60 | **0.006** |
| **V5** | 0.04 | 1.04 (1.01-1.06) | 703 | 0.58 | **0.007** | 0.02 | 1.02 (0.99-1.05) | 350 | 359 | 0.52 | 0.14 |
| **D95** | 0.05 | 1.05 (1.03-1.08) | 704 | 0.61 | **<0.0001** | 0.04 | 1.04 (1-1.07) | 350 | 360 | 0.56 | 0.062 |

| | | | | | | | | | | | |
|---|---|---|---|---|---|---|---|---|---|---|---|
| **D97** | 0.06 | 1.06 (1.03-1.09) | 706 | 0.60 | **0.0001** | 0.04 | 1.04 (0.99-1.08) | 351 | 361 | 0.55 | 0.105 |
| **D98** | 0.06 | 1.06 (1.03-1.09) | 708 | 0.60 | **0.0004** | 0.03 | 1.03 (0.99-1.08) | 352 | 361 | 0.54 | 0.161 |
| **D99** | 0.06 | 1.06 (1.02-1.1) | 710 | 0.59 | **0.001** | 0.03 | 1.04 (0.98-1.09) | 352 | 362 | 0.54 | 0.179 |
| **minGy** | 0.00 | 1 (1-1) | 714 | 0.58 | **0.012** | 0.00 | 1 (1-1) | 353 | 362 | 0.54 | 0.325 |
| **Chemotherapy** | 0.61 | 1.85 (1.06-3.21) | 715 | 0.54 | **0.029** | 0.14 | 1.15 (0.53-2.53) | 353 | 363 | 0.51 | 0.721 |
| **Gender** | 0.60 | 1.83 (0.98-3.41) | 716 | 0.53 | 0.059 | -0.04 | 0.96 (0.42-2.2) | 354 | 363 | 0.50 | 0.926 |
| **postop RT** | 0.52 | 1.68 (1.07-2.65) | 716 | 0.54 | **0.025** | 1.30 | 3.67 (1.96-6.88) | 339 | 348 | 0.63 | **<0.0001** |
| **Age** | -0.01 | 0.99 (0.97-1.01) | 720 | 0.52 | 0.449 | -0.01 | 0.99 (0.96-1.02) | 353 | 362 | 0.53 | 0.369 |
| **pack years** | 0.00 | 1 (0.99-1.01) | 720 | 0.50 | 0.699 | -0.01 | 0.99 (0.98-1.01) | 352 | 362 | 0.52 | 0.251 |
| **Smoking status** | 0.08 | 1.09 (0.62-1.91) | 720 | 0.50 | 0.776 | 1.25 | 3.48 (0.83-14.55) | 349 | 359 | 0.55 | 0.088 |
| **Volume** | 0.00 | 1 (1-1) | 720 | 0.51 | 0.662 | 0.00 | 1 (1-1) | 354 | 363 | 0.52 | 0.817 |

**Appendix 3**

**Figure**. For ORN any grade, frequency plots of variables selected during bootstrapped forward selection (ranked on AIC and with likelihood-ratio test $p<0.01$) in 5000 samples. Position of variable in the model was considered, i.e. forward selection selects per sample most important variable first, and adds variables in subsequent significant importance.

The $D_{30\%}$ was most frequently selected as <u>first</u> variable (blue bar) in 50% of the bootstrap iterations, followed by the $D_{25\%}$ in 23%. The most frequent <u>second</u> selected variable (green bar) was the variable *dental extraction* in 47% of the samples.

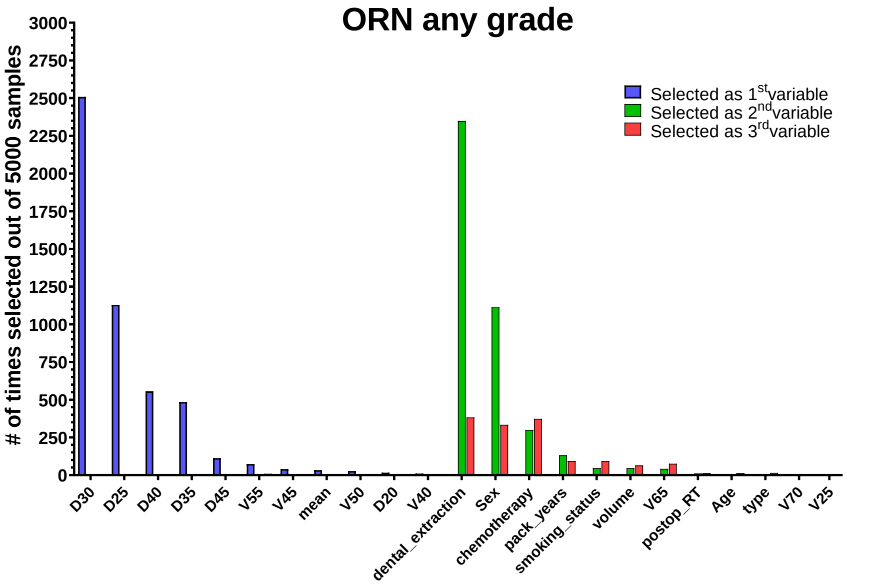

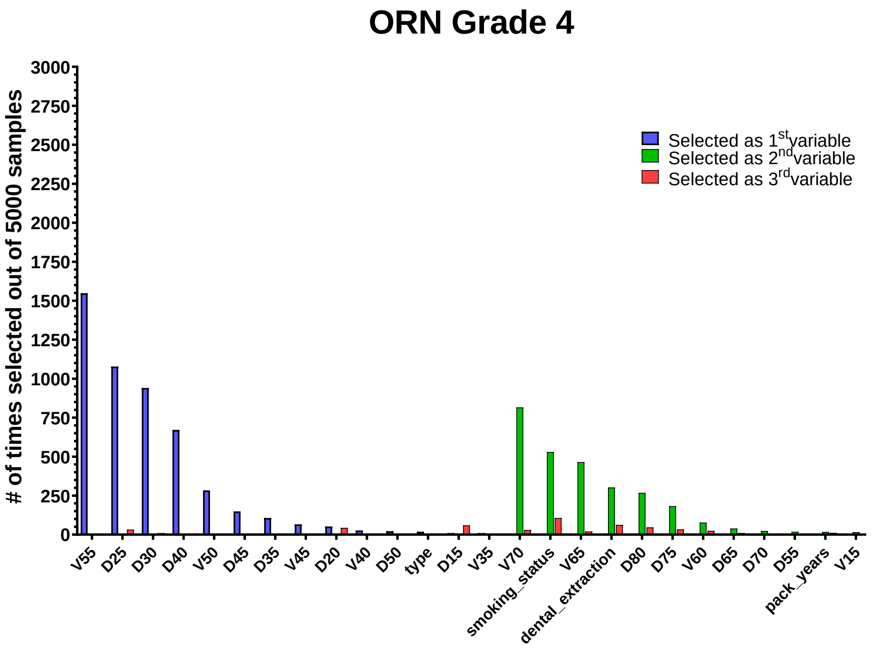

### Appendix 4

**Table**. Performance measures of the full scope of investigated NTCP models for ORN Grade IV

| | Grade IV ORN | | | | | |
|---|---|---|---|---|---|---|
| | D30<br>- | V55<br>- | D30<br>Dental Extractions | D30<br>Smoking status | V55<br>Dental Extractions | V55<br>Smoking status |
| | **Training (n=882)** | | | | | |
| AIC | 306.5 | 306.6 | 304.9 | 302.3 | 303.6 | 302.5 |
| AUC training | 0.80 (0.76-0.85) | 0.82 (0.78-0.87) | 0.81 (0.76-0.86) | 0.81 (0.76-0.86) | 0.82 (0.77-0.87) | 0.82 (0.77-0.87) |
| Nagelkerke $R^2_{training}$ | 0.16 | 0.16 | 0.17 | 0.18 | 0.17 | 0.17 |
| Discrimination slope | 0.05 | 0.06 | 0.06 | 0.06 | 0.07 | 0.07 |
| HL test $X^2$ (p-value) | 18.86 (0.02) | 18.61 (0.02) | 10.82 (0.21) | 11.7 (0.17) | 10.17 (0.25) | 8.58 (0.38) |
| | **Validation (n=377)** | | | | | |
| AUC validation | 0.81 | 0.78 | 0.82 | 0.75 | 0.78 | 0.71 |
| Nagelkerke $R^2_{validation}$ | 0.20 | 0.11 | 0.20 | 0.14 | 0.12 | 0.07 |

**Appendix 5**

Table 1. Performance of final NTCP models (Table 3; $D_{30\%}$ and *Dental extraction*) in sub-cohorts. Where 'Other' are larynx/hypo/nasopharynx/unknown-primary cancer patients, OPC is oropharynx cancer patients

| ORN | Any grade | | | | Grade IV | | |
|---|---|---|---|---|---|---|---|
| | **OPC** | **Oral Cavity** | **Other** | **OPC/Other** | **OPC** | **Oral Cavity** | **OPC/Other** |
| | D30<br>Dental extraction | D30<br>Dental extraction | D30<br>Dental extraction | D30<br>Dental extraction | D30<br>Dental extraction | D30<br>Dental extraction | D30<br>Dental extraction |
| $AUC_{validation}$ | 0.76 | 0.59 | 0.76 | 0.79 | 0.80 | 0.57 | 0.84 |
| Nagelkerke $R^2_{validation}$ | 0.18 | 0.04 | 0.08 | 0.22 | 0.16 | 0.02 | 0.20 |

Table 2. Performance of final NTCP models (Table 3; $D_{30\%}$ and *Dental extraction*) in sub-cohorts of patient treated **definitive** and **post-operative** radiotherapy

| ORN | Any grade | | Grade IV | |
|---|---|---|---|---|
| | **Definitive** | **Post-operative** | **Definitive** | **Post-operative** |
| | D30<br>Dental extraction | D30<br>Dental extraction | D30<br>Dental extraction | D30<br>Dental extraction |
| $AUC_{validation}$ | 0.78 | 0.65 | 0.82 | 0.68 |
| Nagelkerke $R^2_{validation}$ | 0.20 | 0.11 | 0.18 | 0.12 |
